# Self-medication practices and associated factors among COVID-19 recovered patients to prevent future infections: A web-based survey in Bangladesh

**DOI:** 10.1101/2022.05.14.22275075

**Authors:** Md. Safaet Hossain Sujan, Atefehsadat Haghighathoseini, Rafia Tasnim, Md. Saiful Islam, Sarif Mahammad Salauddin, Mohammad Mohiuddin Hasan, Muhammad Ramiz Uddin

## Abstract

**Background:** Human health is largely affected by self-medication in both ways, adversely and favorably, as evidenced by the COVID-19 pandemic. The fear of spreading COVID-19 among health workers and hospital environments has led many Bangladeshi people to practice self-medicate for as a preventive strategy against this disease. Consequently, this practice entails an improper and injudicious use of medicine to cure self-recognized symptoms. To date, the COVID-19 has no effective treatment. The lack of a cure for COVID-19 and the continual progression of the diseases in educational settings induce a substantial population to practice self-medication. Therefore a study of self-medication practices is necessary for the framework of the pandemic. This study aimed to estimate the prevalence and factors associated with self-medication to prevent or manage future COVID-19 infections among recovered COVID-19 patients.

**Methods:** This cross-sectional study was conducted from September 2020 to February 2021 using an e-survey along with 360 participants. Data were collected using a pre-tested self-reported questionnaire. Descriptive statistics and correlations analysis were performed in the study.

**Results:** Among 360 participants, males were 69.7%, and females 30.3%. The prevalence of self-medication is 11%, and monthly family income, residence, education, occupation, and previous history of SM are the associated factors. Among the participants, 29.7% use antibiotics, and 30% use herbal products or drugs as medication.

**Conclusion:** The present study found SMP is moderately prevalent among COVID-19 recovered patients. To minimize the rate of SMP, adequate health care access systems and public education should be introduced, and media & community should be engaged in rational use of medication.

## 1. Introduction

The COVID-19 pandemic has sparked a global lockdown, leaving people with the impression that their only option is self-help, self-care, and self-medication (1). This pandemic caused extreme anxiety and fear among the Bangladeshi people, owing to an increase in confirmed cases as well as a high fatality rate in the South-Asian region, which was exacerbated by the fact that there is no approved vaccine or medication for its treatment. As a result, many people, particularly those who are feeling ill, have turned to the consumption of various drugs, including traditional medicines to treat or prevent a perceived COVID-19 infection without considering the safety and efficacy in the human body (2). Practicing home remedies, herbal foods/medicine as the preventive measures for COVID-19 has also been seen in this country (3). However, lack of swift response, shortage of hospital beds, lack of medical clinics, insufficient test capability (qRT-PCR), (4) the proliferation of unauthenticated treatment procedures placed people in a dilemma to choose professional care.

However, people who are seeking suitable measures to fight the virus, especially people who had already developed some symptoms similar to COVID-19 or recovered from it may get influenced by the COVID-19 related infodemic. Additionally, a substantial number of recovered COVID-19 patients keep practicing it to prevent or manage future infections with the same medicine they used to treat themselves earlier or known to be effective in preventing COVID-19 (5). Consequently, the practice of self-medication among the general public is increasing (6). While in low or middle-income countries, including Bangladesh, the incidence of self-medication may be greater without consultation with trained healthcare professionals (7).

Self-medication (SM), as defined by the WHO, means the selection and use of medicines to treat self-recognized symptoms or disorders without consulting a doctor (8). Also, the use or reuse of previously prescribed or unused medications, direct drug purchases without consultation, and inappropriate medication of over-the-counter (OTC) drugs are often included (9).

SM is a significant global problem affecting developing and developed countries (10). Different studies show that SM is a widespread practice with a worldwide prevalence of 32.5–81.5% (11). SM is widespread but dangerous approach to the management of illnesses that can lead to severe illness, delays in the diagnosis of medical disorders, or enhance the risk of complications (12). The probable reason for extravagance in SM in this country includes past treatment experience with the same symptoms, readily drug availability, halted accessibility to healthcare facilities, socio-cultural or religious values, comparatively high hospital treatment rates, suggestions from friends/family/media (7), ignorance behavior and poverty. In addition, the doctor-patient ratio in Bangladesh is currently as low as it puts the country second from the bottom, among South Asian countries, according to the WHO (13). The socio-economic disparity among people and feeling of helplessness during this pandemic has further increased the unequal access to healthcare since early 2020 (14). Due to frequent symptoms of pain or soreness of the throat, dry cough, severe headache, fever, fatigue, muscle aches, or breathlessness, people started taking the medication without COVID-19 diagnoses or tests (7). Moreover, people diagnosed with COVID-19 are also doing so during the early stages of COVID-19, hoping that the symptoms will not be developed into severe illness. Furthermore, individuals who recovered from it also demonstrated the practice of self-medication to mitigate post-COVID-19 complications.

However, arbitrary use of medicaments can have several risks and side effects, including poisoning, misdiagnosis, medication error (ME) (15), adverse drug reactions (16), endorse drug addiction, promoting drug toxicity, increasing pathogens resistance (12), potentially life-threatening adverse effects, inappropriate dosage, and unreasonable drug use may even lead to death (17). Although, 0.1% of self-medication-related complications will be tons in number, that would be challenging to deal with for the current health care system, which is currently battling against COVID-19.

However, researchers anticipated that the practice of SM among COVID-19 infected and recovered patients, as well as mass people, will be substantially high following the pandemic (1), (Onchonga, Omwoyo, and Nyamamba 2020). Therefore, the present study aimed to investigate the prevalence and associated factors of self-medication practice among COVID-19 recovered patients to prevent or manage future infections of it.

## 2. Methods

### 2.1 Study design, participants, and procedure

A cross-sectional e-survey was conducted from September 2020 to February 2021. A purposive sampling technique was utilized in this study. The study took each participant about 10-12 minutes to complete the entire survey. At first, 402 people took part in the study. After deleting incomplete responses and participants who replied but did not infect with COVID-19, 360 responses were included in the final analysis. A self-reported questionnaire developed in Bangla (the participant’s native language) was utilized to collect data during the study. Following the completion of all questions in the Google Form, a shareable link was provided. The survey link was shared on several community-based online forums to elicit replies from individuals who had recovered from COVID-19.

A pilot test was conducted with 35 participants within the same population (target group) to check the questionnaire’s acceptability, validity, and transparency before moving on to the next phase. Minor changes were made to the questionnaire after the pilot testing. Those questions with responses were not included in the final analysis. An informed consent statement outlining the purpose and process and the right to refuse participation in the study was attached to the first page of the questionnaire. Individuals were requested to obtain written informed permission before being asked, “Are you willing to take part in this study voluntarily and spontaneously?” A blank survey form was automatically submitted if the person replied ‘no’. If the person replied *‘Yes,’* he/she was allowed access to the full form of the survey. In the study, the exclusion criteria for participation were individuals below 18 years old, with no COVID-19 infection confirmation, and severe psychological problems (as this can cause memory biases).

### 2.2 Measures

#### 2.2.1 Socio-demographic measures and determinants of self-medication

In the present study, socio-demographic information was collected by asking questions about sex (male/female), age (younger adults >18-39 years, middle-aged >40 years), residence (rural/urban), occupation (unemployed/employed), education (tertiary/equal or below secondary), smoking habit (yes/no). Depending on their monthly total family income in Bangladeshi Taka (BDT), socioeconomic status (SES) was divided into three classes: lower SES [15,000 BDT], medium SES [15,000-30,000 BDT], and upper SES [> 30,000 BDT](19).

#### 2.2.2 Self-medication related questions

Measures related to self-medication were obtained from previous studies (2), (20) as well as by asking questions as follows: *“What was your* Action after identifying the symptoms of COVID-19*?”* (Waiting for cure/ Consulting with doctor/ Self-medication), *“Do you have* health care professionals in family members*?”* (yes/no), *“Do you have* the Previous history of self-medication*?”* (yes/no), *“Do you think self-medication is harmful?”* (yes/no), “Prescription of medication given by? “ (Myself/Physician/Specialist), “Knowledge on self-medication? “ (Sufficient/Insufficient), *“ Self-medicating medicine?”* (Antibiotic/ Doxycycline/ dexamethasone/ Ivermectin/Favipiravir), *“Do you use* Herbal products/drugs for treatment*?”* (yes/no), *“What was your reason to practice self-medication ?”* (Familiar medicine/ Mild symptoms/ To save time/Readily available/ To get the quick result/Financial crisis).

### 2.3 Statistical analysis

Microsoft Excel 2019 and IBM SPSS Statistics version 25 were used to analyze the data. Microsoft Excel is used to clean, code, and sort data. SPSS software was used to perform descriptive statistics (e.g., frequencies, percentages, means, standard deviations, and so on). For the regression analysis, a p-value of less than 0.05 was considered significant.

### 2.4 Ethical Approval

The study was conducted following the Institutional Research Ethics and Human Involvement Guidelines (Helsinki declaration). The Ethical Review Committee gave their approval for the study. Formal ethical approval was granted from Ethical Review Committee, Jahangirnagar University [BBEC, JU/M/ COVID-19(7)4]. The purpose of this study, as well as i) the methods and objectives of the current investigation, ii) data confidentiality and anonymity, and iii) the flexibility to withdraw response from the study at any moment, were all documented in the first phase of the questionnaire.

## 3. Results

### 3.1 Socio-demographic characteristics

A total of 360 participants were included in the final analysis. Among them, 69.7% were male and 30.3% were female. The majority of the participants were younger adults (>18-39 years) (58.1%), and middle-aged (>40). The majority of the participants (84.7%) are unemployed, about 74.4% of the participants completed secondary level education, and a, among the participants, substantial proportion (60.3%) have smoking habits. Most of the participants were from middle socioeconomic status (41.9%) and resided in urban areas (71.9%). Among the participants, 32.5% got the COVID-19 test after identifying the symptoms of COVID-19, 31.7% consulted with doctors, and only 23.9% of the participants waited for a cure by itself. The majority of the participants (61.9%) had no family member in the health care profession. Half of the participants (51.1%) had never self-medicated before, and the majority of the people (71.7%) believe that self-medication is harmful. The majority of the respondents (30.6%) took paracetamol, and antibiotic (29.7%) to treat their symptoms, and 70% of the respondents didn’t take any herbal products/drugs for their treatment.

### 3.2 Factors affecting self-medication

Self-medication was found to be strongly associated with the participants’ residence in this study. People living in the rural area (17.8%) score a significantly higher rate of self-medication than people in urban areas, and only 9.7% of urban participants practiced SM. But the age and sex were not significantly associated with self-medication. The previous history of self-medication was found significantly associated with self-medication. Respondents with prior self-medication history (21.0%) score higher than respondents with no self-medication history. Moreover, 90.3% of the respondents believe that self-medication is harmful to health and didn’t self-medicate, and 17.6% of the respondents believe that self-medication is not harmful to health and continued self-medicating themselves. About 83.6% of the respondents practice self-medication by their own decision, and a large proportion (58.6%) of participants have sufficient self-medication related knowledge. The main causes of self-medication was the familiar medicine for used for previous diseases (35.3%). Among the participants (15.7%) used herbal medicine as alternative medicine, and (35.3%) practice it as the medicine was familiar to them due to previous use, and (12.5%) use it to get quick results.

**Table 1:**
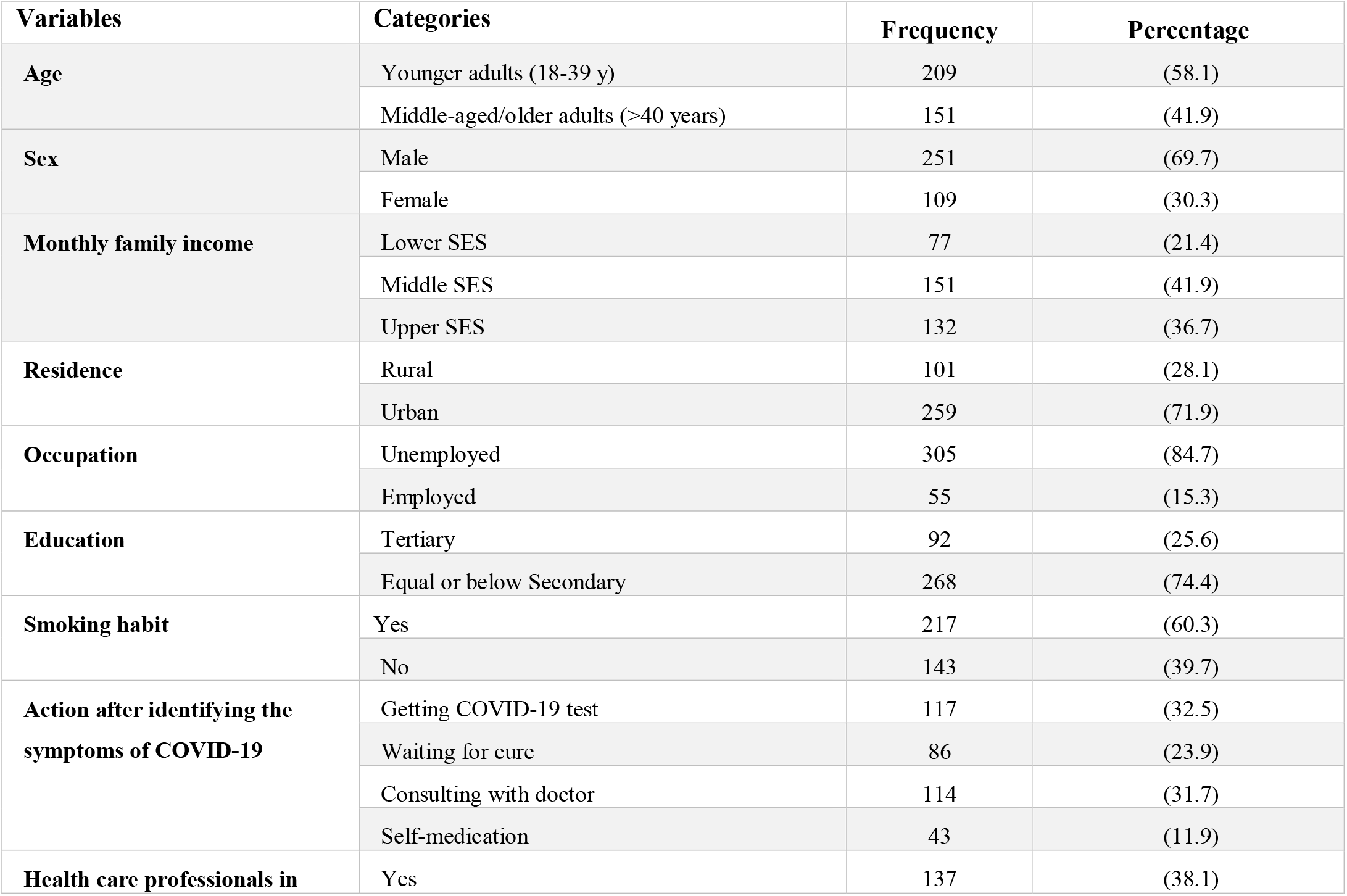
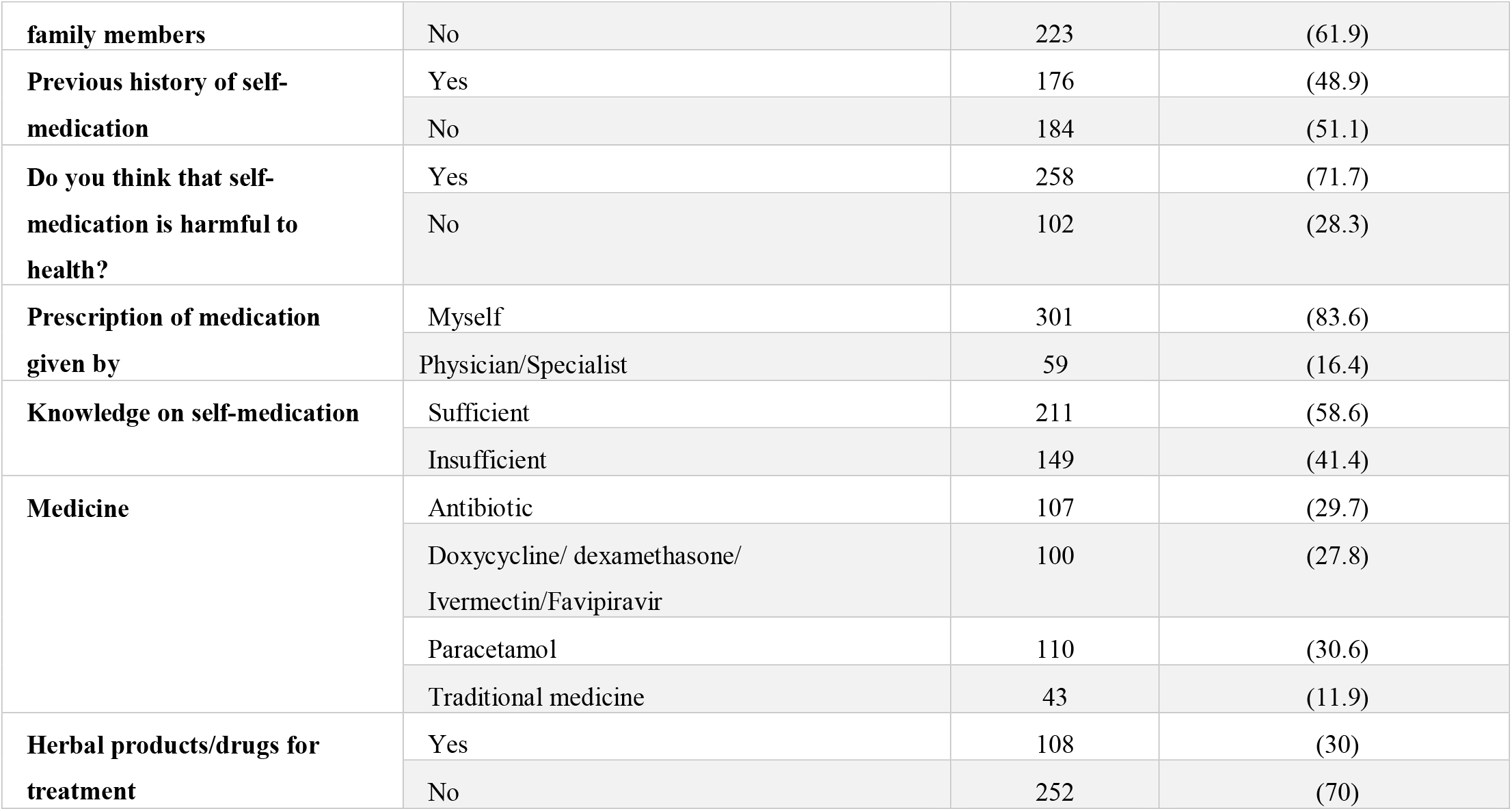
General characteristics of participants.

**Figure:**
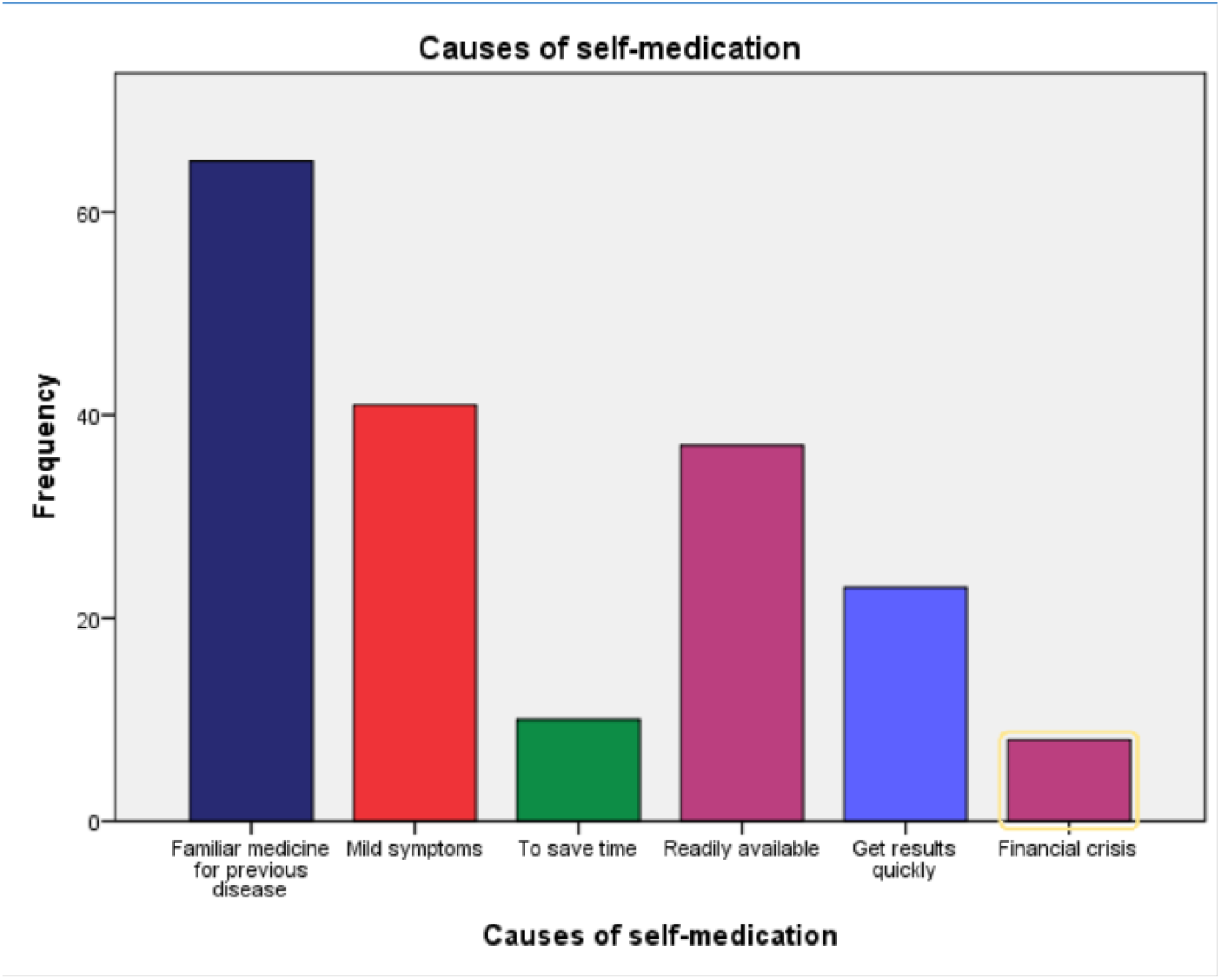
Probable causes of self-medication.

**Table 2:** Association between self-medication and other variables.

| Variables | Categories | Self-medication |  |  |  | *p*-value |
| --- | --- | --- | --- | --- | --- | --- |
|  |  | *No* |  | *Yes* |  |  |
|  |  | **n** | **(%)** | **n** | **(%)** |  |
| Age | Younger adults | 181 | (86.6) | 28 | (13.4) | 0.317 |
|  | Middle-aged/older adults | 136 | (90.1) | 15 | (9.9) |  |
| Sex | Male | 219 | (87.3) | 32 | (12.7) | 0.475 |
|  | Female | 98 | (89.9) | 11 | (10.1) |  |
| Monthly family income | Lower SES | 68 | (88.3) | 9 | (11.7) | **0.057** |
|  | Middle SES | 130 | (86.1) | 21 | (13.9) |  |
|  | Upper SES | 119 | (90.2) | 13 | (9.8) |  |
| **Residence** | Rural | 83 | (82.2) | 18 | (17.8) | **0.032** |
|  | Urban | 234 | (90.3) | 25 | (9.7) |  |
| **Occupation** | Unemployed | 267 | (87.5) | 38 | (12.5) | **0.047** |
|  | Employed | 50 | (90.9) | 5 | (9.1) |  |
| **Education** | Tertiary | 76 | (82.6) | 16 | (17.4) | **0.052** |
|  | Equal or below Secondary | 241 | (89.9) | 27 | (10.1) |  |
| **Smoking habit** | Yes | 189 | (87.1) | 28 | (12.9) | 0.490 |
|  | No | 128 | (89.5) | 15 | (10.5) |  |
| **Health care professionals in family members** | Yes | 124 | (90.5) | 13 | (9.5) | 0.260 |
|  | No | 193 | (86.5) | 30 | (13.5) |  |
| **Previous history of self-medication** | Yes | 139 | (79.0) | 37 | (21.0) | **<0.001** |
|  | No | 178 | (96.7) | 6 | (3.3) |  |
| **Do you think that self-medication is harmful to health?** | Yes | 233 | (90.3) | 25 | (9.7) | **0.036** |
|  | No | 84 | (82.4) | 18 | (17.6) |  |
| **Prescription of medication given by** | Myself | 264 | (87.7) | 37 | (12.3) | .646 |
|  | Physician/Specialist | 53 | (89.8) | 6 | (10.2) |  |
| **Knowledge on self-medication** | Sufficient | 184 | (87.2) | 27 | (12.8) | **0.055** |
|  | Insufficient | 133 | (89.3) | 16 | (10.7) |  |
| **Medicine** | Antibiotic | 93 | (86.9) | 14 | (13.1) | **0.022** |
|  | Doxycycline/ dexamethasone/ Ivermectin/Favipiravir | 93 | (93.0) | 7 | (7.0) |  |
|  | Paracetamol | 96 | (87.3) | 14 | (12.7) |  |
|  | Traditional medicine | 35 | (81.4) | 8 | (18.6) |  |
| **Herbal products/drugs for treatment** | Yes | 91 | (84.3) | 17 | (15.7) | **0.014** |
|  | No | 226 | (89.7) | 26 | (10.3) |  |

## 4. Discussion

The purpose of this study was to estimate the prevalence, causes, and determinants of self-medication practices for COVID-19 prevention and/or treatment in Bangladesh. Though the world health organization (WHO) recommends self-medication for minor illnesses (2), our study looked into self-medication for perceived COVID-19 prevention/treatment. To the best of our knowledge, this is the first study on the objective of COVID-19 related self-medication practices among recovered COVID-19 patients, though there have been previous studies on self-medication practices other than COVID-19 in Bangladesh and elsewhere.

People have long been concerned with their health and have practiced self-medication since ancient times. Although there are numerous advantages and disadvantages of practicing self-medication, it all relies on who uses it and how it is utilized for self-treatment. While this is undeniable, self-medication without consulting with certified health professionals may be more common in low- and middle-income countries. SMP is common in most sections of Bangladesh, regardless of socioeconomic class or educational attainment (7). SM may be a low-cost alternative for people in impoverished countries like Bangladesh to avoid the high expense of clinical services and many medications available over the counter (OTC) without a prescription (21). However, to the best of the author’s knowledge, there is no current evidence on the prevalence of SMP among COVID-19 recovered patients in Bangladesh. In addition, there is a dearth of knowledge globally regarding this issue.

However, in this study, we focused on self-medication practices among COVID-19 recovered patients in Bangladesh to estimate the prevalence of self-medication practices and the factors associated with them. This study found 11% self-medication practice among COVID-19 recovered patients, which is much less than a couple of studies previously conducted in Bangladesh among general populations (12), (7), among rural and urban people in India (22), (23), among general populations in Pakistan (24), in general resident in China (25), among health science students in Iran (26), and among school students parents in Italy (27). The prevalence is less than those studies may be because of different study settings, time, and target groups. Additionally, the present study conducted among the COVID-19 recovered patients who practiced SM to prevent or manage future infections of COVID-19, this could be another significant cause of low prevalence. As the study was conducted among COVID-19 recovered patients, therefore the patients are exceptionally concerned regarding the disease and many are afraid of practicing SM. Moreover, there was no effective treatment discovered to be practiced which could be another reason.

However, according to this study, self-medication practice (SMP) has been found significantly associated with a person’s residence, and the prevalence of SMP is higher among people living in the rural area than people living in urban areas. In contrast to a study in Ethiopia that revealed that living in a city was strongly associated with self-medication (28). SMP was shown to be more common among those living in large cities in a similar study in Spain (29). Another study in rural parts of Portugal found a 21.5% prevalence of SMP among rural people (30). This may be due to the lower literacy level among individuals living in rural areas. A study found Bangladesh, as one of the world’s poorest countries, has extremely low educational attainment, especially among women (31). Only 9% of boys and 5% of girls finish high school (31) in rural areas. This may be a cause of self-medication as a previous study found a significant association between a father’s education and self-medication (32). People living in rural areas might have to travel long distances and pay a lot of money to visit a doctor (33), The majority of Bangladesh’s rural areas lack a sufficient number of dispensaries, and those that do exist are ineffective due to a shortage of medical personnel and medications during the pandemic. SMP is found among people in lower socioeconomic status in this present study which is in line with a previous study in Tamil Nadu (34). According to this study, unemployment was found to be significantly associated with self-medication. In contrast to a study in Tamil Nadu, where employed participants had a higher rate of self-medication (34). Many residents in rural districts rely on daily wages to supplement their income because they commute to the city every day but due to COVID-19 lockdown, thousands of people lost their jobs and suffered from financial crisis to consult a doctor because of the high cost of medical consultations (35,36), these factors may also lead low socioeconomic status people into self-medication.

Tertiary education or university graduates has been found to be significantly associated with self-medication. According to a prior study conducted on adolescents, those with lower medical literacy are more prone to participate in inappropriate self-medication (37). Another reason could be that educated people are more confident about the effects of medicines and practice it randomly as they believe this would be less harmful. Nonetheless, people who are sick frequently treat themselves, which is likely owing to the human survival instinct. People act on their health every day, all across the world, without consulting competent medical professionals. They make it a habit and a culture to take care of themselves.

However, the present study found that SMP was significantly associated with the previous history of self-medication and it is higher among people who suffered from the same illness previously. According to a previous study among Bangladeshi students, the largest proportion (38%) of students learned self-medication from an old doctor’s prescription to treat present illness (38). Prior studies in the same country found that previous prescriptions were the most important source of information and the most important factor of self-medication (39) and the pre-experience was one of the main causes for self-medication of antibiotics (40). The findings that the majority of respondents got information regarding self-medication from previously prescribed medicines by doctors were consistent with research among Brazilian universities (41). Inappropriate SMP can result from misplaced confidence, and individuals may be exposed to all of the risks involved with self-medication. According to studies from the United States, Asia, and Europe, 22-70% of parents have misconceptions regarding the proper usage and efficacy of antibiotics (40). However, other factors, such as the opinions of family members, friends, neighbors, and advertisements, may have impacted the responders (39). The fact that such a large percentage of people self-medicate on their initiative highlights the necessity for health care practitioners to launch an intensive public awareness campaign about the dangers of self-medication. A further longitudinal study is needed to properly determine the association between self-medication and history of illness.

In addition, in this study, we found that majority of the participants were aware of the dangerous effects of self-medication. According to a previous study conducted in the UAE, the majority of respondents (64.1 %) considered self-medication was safe (39). Another survey found that 47% of participants believe self-medication is a form of self-care that should be encouraged (22), which aligns with other studies from Ethiopia and Karachi (24,42). According to a telephone survey in the USA, 58 % of people were unaware of the potential health risks associated with antibiotic use (43).

SMP has found to be significantly associated with knowledge of an individual given by prescription of medication. In contrast to a prior study among adolescents, participants with lower medication awareness were more prone to engage in inappropriate self-medication (37).

Though, there are several advantages to appropriate self-medication, including rising patient access to medication and relief, the patient’s prominent participation in their health care, improved use of physicians’ and pharmacists’ skills, and lowered (or at least optimized) government burden due to health expenditure in this pandemic associated to the treatment of minor health conditions (44). However, Self-medication, on the other hand, is far from being a perfectly safe activity, especially in the event of irresponsible self-medication. Erroneous self-diagnosis, postponement in seeking medical advice when needed, occasional but severe adverse reactions, dangerous drug interactions, incorrect method of administration, wrong medication, inappropriate preference of treatment, masking of severe disease, and the risk of dependence and abuse are all potential risks of self-medication (44). Government officials should keep a careful eye on the rules and regulations governing the sale of pharmaceuticals from pharmacies. To adequately educate people about the adverse effects of self-medication, a holistic approach should be used, including sufficient awareness and education on self-medication and stricter regulations on pharmaceutical advertising. These programs might be conducted online due to pandemic-related restrictions.

The World Health Organization (WHO) and epidemiologists from many countries have anticipated that the COVID-19 pandemic will last for years and have a significant socioeconomic and psychosocial influence on people’s lifestyles and behavior. Without appropriate scientific proof, a variety of medicines were utilized to treat respiratory and COVID-19-related symptoms. Antibiotics and paracetamol were the most commonly used pharmaceuticals, while Doxycycline, dexamethasone, Ivermectin, Favipiravir, and even traditional treatments were widely used. They were used as COVID-19 preventives to treat suspected symptoms, and even after a positive COVID-19 diagnosis. This study may provide information to health educators, planners, and other health professionals that will aid in the reduction of SM and the promotion of positive and responsible medicine. A comprehensive nationwide survey and surveillance of mass population self-medication should be conducted to safeguard them from potential hazards, overuse shortages, and unjustified financial engagement during the COVID19 outbreak.

### Limitations

There is no study outside constraints, and this study has some limitations as well. Firstly, the study was cross-sectional in its nature, therefore, the causality of factors could not be established. In this regard, a longitudinal study is required to better understand the practice of self-medication among COVID-19 recovered patients. Secondly, the study employed an online-based self-reporting method, which could have been influenced by a variety of biases, including perceived benefits and episodic memory biases. Thirdly, due to the inflexibility of reaching out to persons who had recovered from COVID-19 and their refusal to engage in this study freely, the study enrolled only a few participants.

## 5. Conclusion

SMP has become a significant public health concern in Bangladesh, particularly during the COVID-19 pandemic. This study revealed a considerable number of COVID-19 recovered patients who practiced self-medication to prevent or manage the future occurrence of COVID-19. One in every eleven participants who recovered from COVID-19 practice self-medication. It is necessary to maintain constant knowledge and sensitization about the dangers of self-medication. Our findings should be viewed with caution and should not be construed as a piece of advice to self-medicate or to use these medications in the hopes of improving symptomatology. Before taking any medication, always seek medical advice and consultation. We anticipate that these findings will help healthcare policymakers make better decisions about how to improve pharmaceutical care and save lives.

## Data Availability

All data produced in the present study are available upon reasonable request to the authors

## Author’s Contribution

Conceptualization: MSHS, AH, RT, MSI, Methodology: MSHS, MSI, Formal Analysis: MSHS, MSI, Validation: MSHS, AH, RT, MSI, SMS, MMH, MRU, Investigation & Writing Original Draft: MSHS, AH, RT, MSI, SMS, Data Curation: MSHS, Review & Editing: SMS, MMH, MRU. All authors have read and agreed to the published this version of the manuscript.

## Acknowledgements

We thank all the participants who took part in the study. We also acknowledge the efforts of all research assistants that helped in data collection for the study.

## Data Availability

The data that support the fndings of the current study are available from the corresponding author upon reasonable request.

## Consent for Publication

Not applicable

## References

1. Quispe-Cañari JF, Fidel-Rosales E, Manrique D, Mascaró-Zan J, Huamán-Castillón KM, Chamorro–Espinoza SE, et al. Self-medication practices during the COVID-19 pandemic among the adult population in Peru: A cross-sectional survey. Saudi Pharm J. 2021;29(1):1–11.

2. Wegbom AI, Edet CK, Raimi O, Fagbamigbe AF, Kiri VA. Self-Medication Practices and Associated Factors in the Prevention and / or Treatment of COVID-19 Virus_J: A Population-Based Survey in Nigeria. 2021;9(June):1–9.

3. Ahmed I, Hasan M, Akter R, Sarkar BK, Rahman M, Sarker MS, et al. Behavioral preventive measures and the use of medicines and herbal products among the public in response to Covid-19 in Bangladesh: A cross-sectional study. PLoS One. 2020;15(12):e0243706.

4. Islam S, Islam R, Mannan F, Rahman S, Islam T. COVID-19 pandemic: An analysis of the healthcare, social and economic challenges in Bangladesh. Prog Disaster Sci. 2020;8:100135.

5. Quincho-Lopez A, Benites-Ibarra CA, Hilario-Gomez MM, Quijano-Escate R, Taype-Rondan A. Self-medication practices to prevent or manage COVID-19: A systematic review. PLoS One [Internet]. 2021;16(November):1–12. Available from: http://dx.doi.org/10.1371/journal.pone.0259317

6. Mallhi TH, Khan YH, Alotaibi NH, Alzarea AI, Alanazi AS, Qasim S, et al. Drug repurposing for COVID-19: a potential threat of self-medication and controlling measures. Postgrad Med J. 2021;97(1153):742–3.

7. Nasir M, Chowdhury ASMS, Zahan T. Self-medication during COVID-19 outbreak: a cross sectional online survey in Dhaka city. Int J Basic Clin Pharmacol. 2020;9(9):1325.

8. WHO. Guidelines for the regulatory assessment of medicinal products for use in self-medication. 2000.

9. Eticha T, Mesfin K. Self-medication practices in Mekelle, Ethiopia. PLoS One. 2014;9(5):e97464.

10. Malik M, Tahir MJ, Jabbar R, Ahmed A, Hussain R. Self-medication during Covid-19 pandemic: challenges and opportunities. Drugs Ther Perspect. 2020;36(12):565–7.

11. Kassie AD, Bifftu BB, Mekonnen HS. Self-medication practice and associated factors among adult household members in Meket district, Northeast Ethiopia, 2017. BMC Pharmacol Toxicol. 2018;19(1):15.

12. Moonajilin MS, Mamun MA, Rahman ME, Mahmud MF, Al Mamun AHMS, Rana MS, et al. Prevalence and drivers of self-medication practices among savar residents in Bangladesh: A cross-sectional study. Risk Manag Healthc Policy. 2020;13:743–52.

13. WHO. Bangladesh. 2013.

14. Kretchy IA, Asiedu-Danso M, Kretchy J-P. Medication management and adherence during the COVID-19 pandemic: perspectives and experiences from low-and middle-income countries. Res Soc Adm Pharm. 2020;17(1):2023–6.

15. Mira JJ, Lorenzo S, Guilabert M, Navarro I, Pérez-Jover V. A systematic review of patient medication error on self-administering medication at home. Expert Opin Drug Saf. 2015;14(6):815–38.

16. Berreni A, Montastruc F, Bondon□Guitton E, Rousseau V, Abadie D, Durrieu G, et al. Adverse drug reactions to self_Jmedication: a study in a pharmacovigilance database. Fundam Clin Pharmacol. 2015;29(5):517–20.

17. Khatony A, Soroush A, Andayeshgar B, Abdi A. Nursing students’ perceived consequences of self-medication: a qualitative study. BMC Nurs. 2020;19(1):1–7.

18. Onchonga D, Omwoyo J, Nyamamba D. Assessing the prevalence of self-medication among healthcare workers before and during the 2019 SARS-CoV-2 (COVID-19) pandemic in Kenya. Saudi Pharm J [Internet]. 2020;28(10):1149–54. Available from: https://doi.org/10.1016/j.jsps.2020.08.003

19. Islam MS, Sujan MSH, Tasnim R, Ferdous MZ, Masud JHB, Kundu S, et al. Problematic internet use among young and adult population in Bangladesh: Correlates with lifestyle and online activities during the COVID-19 pandemic. Addict Behav Reports [Internet]. 2020;12(November):100311. Available from: https://doi.org/10.1016/j.abrep.2020.100311

20. Okoye OC, Adejumo OA, Opadeyi AO, Madubuko CR, Ntaji M, Okonkwo KC, et al. Self medication practices and its determinants in health care professionals during the coronavirus disease-2019 pandemic: cross-sectional study. Int J Clin Pharm [Internet]. 2022;(0123456789). Available from: https://doi.org/10.1007/s11096-021-01374-4

21. Hussain S, Malik F, Hameed A, Ahmad S, Riaz H. Exploring health seeking behavior, medicine use and self medication in urban and rural Pakistan. South Med Rev. 2010;3(2):32–5.

22. Kumar N, Kanchan T, Unnikrishnan B, Rekha T, Mithra P, Kulkarni V, et al. Perceptions and practices of self-medication among medical students in coastal South India. PLoS One. 2013;8(8):e72247.

23. I. H, S. S, Jayan M, Hussain C. Prevalence of self-medication practices and its associated factors in rural Bengaluru, Karnataka, India. Int J Community Med Public Heal. 2016;3(6):1481–6.

24. Zafar SN, Syed R, Waqar S, Zubairi AJ, Vaqar T, Shaikh M, et al. Self-medication amongst university students of Karachi: prevalence, knowledge and attitudes. J Pak Med Assoc. 2008;58(4):214.

25. Lei X, Jiang H, Liu C, Ferrier A, Mugavin J. Self-medication practice and associated factors among residents in Wuhan, China. Int J Environ Res Public Health. 2018;15(1).

26. Abdi A, Faraji A, Dehghan F, Khatony A. Prevalence of self-medication practice among health sciences students in Kermanshah, Iran. BMC Pharmacol Toxicol. 2018;19(1):1–7.

27. Garofalo L, Di Giuseppe G, Angelillo IF. Self-medication practices among parents in italy. Biomed Res Int. 2015;2015.

28. Mekuria AB, Birru EM, Tesfa MT, Geta M, Kifle ZD, Amare T. Prevalence and predictors of self-medication practice among teachers’ education training college students in Amhara region, Ethiopia: a cross-sectional study. Front Pharmacol. 2020;11.

29. Figueiras A, Caamano F, Gestal-Otero JJ. Sociodemographic factors related to self-medication in Spain. Eur J Epidemiol. 2000;16(1):19–26.

30. de Melo MN, Madureira B, Ferreira APN, Mendes Z, da Costa Miranda A, Martins AP. Prevalence of self-medication in rural areas of Portugal. Pharm World Sci. 2006;28(1):19– 25.

31. Khandker SR, Samad HA. Education Achievements and School Efficiency in Rural Bangladesh. Bangladesh Dev Stud. 1995;1–28.

32. Lukovic JA, Miletic V, Pekmezovic T, Trajkovic G, Ratkovic N, Aleksic D, et al. Self-medication practices and risk factors for self-medication among medical students in Belgrade, Serbia. PLoS One. 2014;9(12):e114644.

33. Keya KT, Rob U, Rahman MM, Bajracharya A, Bellows B. Distance, transportation cost, and mode of transport in the utilization of facility-based maternity services: evidence from rural Bangladesh. Int Q Community Health Educ. 2014;35(1):37–51.

34. Aljin V, Stephen T, Krishnakumar J. PREVALENCE AND DETERMINANTS OF SELF-MEDICATION PRACTICE IN AN URBAN AREA OF KANCHEEPURAM DISTRICT, TAMILNADU. Ann Rom Soc Cell Biol. 2021;2811–21.

35. Posel D, Oyenubi A, Kollamparambil U. Job loss and mental health during the COVID-19 lockdown: Evidence from South Africa. PLoS One. 2021;16(3):e0249352.

36. Fainzang S. Managing medicinal risks in self-medication. Drug Saf. 2014;37(5):333–42.

37. Lee C-H, Chang F-C, Hsu S-D, Chi H-Y, Huang L-J, Yeh M-K. Inappropriate self-medication among adolescents and its association with lower medication literacy and substance use. PLoS One. 2017;12(12):e0189199.

38. Islam B, Hossain MA. Prevalence of self medication practice among students of a medical college. Bangladesh Med J Khulna. 2019;52(1–2):21–4.

39. Seam M, Reza O, Bhatta R, Saha BL, Das A, Hossain M, et al. Assessing the perceptions and practice of self-medication among Bangladeshi undergraduate pharmacy students. Pharmacy. 2018;6(1):6.

40. Biswas M, Roy MN, Manik MIN, Hossain MS, Tapu SMTA, Moniruzzaman M, et al. Self medicated antibiotics in Bangladesh: a cross-sectional health survey conducted in the Rajshahi City. BMC Public Health. 2014;14(1):1–7.

41. Pereira CM, Alves VF, Gasparetto PF, Carneiro DS, de Carvalho D de GR, Valoz FEF. Self-medication in health students from two Brazilian universities. RSBO Rev Sul-Brasileira Odontol. 2012;9(4):361–7.

42. Gutema GB, Gadisa DA, Kidanemariam ZA, Berhe DF, Berhe AH, Hadera MG, et al. Self-medication practices among health sciences students: the case of Mekelle University. J Appl Pharm Sci. 2011;1(10):183.

43. Eng J Vanden, Marcus R, Hadler JL, Imhoff B, Vugia DJ, Cieslak PR, et al. Consumer attitudes and use of antibiotics. Emerg Infect Dis. 2003;9(9):1128.

44. Ruiz ME. Risks of self-medication practices. Curr Drug Saf. 2010;5(4):315–23.

